# Effect of Fathers in Preemie Prep for Parents (P3) Program on Couple’s Preterm Birth Preparedness

**DOI:** 10.1101/2024.09.11.24313503

**Authors:** Mir A. Basir, Siobhan M. McDonnell, Ruta Brazauskas, U. Olivia Kim, S. Iqbal Ahamed, Jennifer J. McIntosh, Kris Pizur-Barnekow, Michael B. Pitt, Abbey Kruper, Steven R. Leuthner, Kathryn E. Flynn

**Affiliations:** Department of Pediatrics, Medical College of Wisconsin, Milwaukee, USA; Division of Biostatistics, Medical College of Wisconsin, Milwaukee, USA; Department of Pediatrics, Northwestern University Feinberg School of Medicine, Chicago, USA; Department of Computer Science, Marquette University, Milwaukee, USA; Department of Obstetrics & Gynecology, Medical College of Wisconsin, Milwaukee, USA; Families First LLC, Eagle River, USA; Department of Pediatrics, University of Minnesota Masonic Children’s Hospital, Minneapolis, USA; Department of Medicine, Medical College of Wisconsin, Milwaukee, USA

**Author notes:** **Corresponding author at**: Mir A. Basir, Department of Pediatrics, Medical College of Wisconsin, 8701 W. Watertown Plank Rd, Milwaukee, WI 53226, USA. **Funding:** This work was support by Eunice Kennedy Shriver National Institute of Child Health and Human Development (NICHD) [grant number R21 HD092664, 2019]. **Clinical Trial Registration**: ClinicalTrials.gov, NCT04093492, https://clinicaltrials.gov/study/NCT04093492. **Abbreviations**: Eunice Kennedy Shriver National Institute of Child Health and Human Development (NICHD); gestational age (GA); neonatal intensive care unit (NICU); American College of Obstetricians and Gynecologists (ACOG); Preemie Prep for Parents (P3); Patient-Reported Outcomes Measurement Information System (PROMIS); Computerized Adaptive Testing (CAT); Parent Prematurity Knowledge Questionnaire (PPKQ).

## Abstract

**Objective:** Evaluate the effect of fathers’ participation in the Preemie Prep for Parents (P3) program on maternal learning and fathers’ preterm birth knowledge.

**Methods:** Mothers with preterm birth predisposing medical condition(s) enrolled with or without the baby’s father and were randomized to the P3 intervention (text-messages linking to animated videos) or control (patient education webpages). Parent Prematurity Knowledge Questionnaire assessed knowledge, including unmarried fathers’ legal neonatal decision-making ability.

**Results:** 104 mothers reported living with the baby’s father; 50 participated with the father and 54 participated alone. In the P3 group, mothers participating with the father (n=33) had greater knowledge than mothers participating alone (n=21), 85% correct responses vs. 76%, *p*=0.033. However, there was no difference in knowledge among the control mothers, 67% vs. 60%, *p*=0.068. P3 fathers (n=33) knowledge scores were not different than control fathers (n=17), 77% vs. 68%, *p*=0.054. Parents who viewed the video on fathers’ rights (n=58) were more likely than those who did not (n=96) to know unmarried fathers’ legal inability to decide neonatal treatments, 84% vs. 41%, *p*<0.001.

**Conclusions:** Among opposite-sex cohabitating couples, fathers’ participation in the P3 program enhanced maternal learning.

**Practice Implications:** The P3 program’s potential to educate fathers may benefit high-risk pregnancies.

## 1. Introduction

Fathers’ involvement in pregnancy is associated with improved outcomes for maternal and neonatal health, including fewer preterm births (1,2). However, fathers experience greater barriers than mothers to prenatal preterm birth education; such education is typically provided during clinic visits arranged around the mother’s schedule. The lack of inclusion is consequential, with fathers reporting that insufficient knowledge contributes to their hesitancy to actively participate in pregnancy care (3). Similarly, though the father’s support for breastfeeding is a strong predictor of mother’s behavior (4,5), fathers report they do not receive sufficient information on breastfeeding (6,7). Both fathers and mothers report neglect of the father by healthcare providers and exclusion from informational provision (8). To further distance fathers from pregnancy and infant care, there are often legal restrictions on unmarried fathers’ ability to participate in decision making (9). In states such as Wisconsin, fathers of the baby are not legally considered parents at birth unless they are married to the baby’s mother (10).

In our clinical trial of the Preemie Prep for Parents (P3) smartphone preterm birth video- education program, pregnant mothers randomized to the P3 intervention had more knowledge of core competencies, including available neonatal resuscitation options (11), and were more prepared to make decisions that impact maternal and infant health, without experiencing increased anxiety, compared to participants assigned to the control arm (12). The P3 trial also enrolled fathers; this manuscript presents the effect of the P3 program on participating fathers and the influence of father’s participation on the mother.

## 2. Methods

The study was approved by the Medical College of Wisconsin Institutional Review Board and conducted at an academic medical center near Milwaukee, Wisconsin. This manuscript follows CONSORT reporting guidelines.

Pregnant mothers with medical conditions that predispose them to preterm birth were recruited from the high-risk obstetric clinic between February 2020 and April 2021 when they were between 16 weeks 0 days and 21 weeks 6 days gestational age (GA). Those unable to speak English and pregnancies with significant anomalies were excluded. Participants were randomized 1:1 to the P3 program or control education. Randomization sequence was generated using R package blockrand and implemented through Research Electronic Data Capture (REDCap) (13).

At enrollment, pregnant mothers were given the opportunity to provide information for a person who would support them through the pregnancy. This support person could be, but was not required to be, the father of the baby. The support person was contacted by phone and invited to participate. Participants provided written consent. To prevent contamination, support persons were assigned to the same study group that the mother was randomized to. The trial ended when the predetermined sample size of 120 pregnant mothers (12) was reached.

The P3 program included 51 animated videos, each 2-3 minutes long and conveying information recommended (14) for parents at risk of preterm birth. Content includes information on signs of preterm labor, risk appropriate birth hospitals, and neonatal resuscitation decisions. Three of the videos focused on mother’s milk, including the unique benefits for preterm infants and how breastfeeding can be facilitated in the neonatal intensive care unit. One video focused on the involvement and role of a support person in pregnancy and another video focused on the legal standing of fathers to make neonatal treatment decisions at birth depending on marital status. Links to videos were delivered by text messages to participants’ smartphones from as early as 18 weeks GA until either 34 weeks GA or preterm delivery. Pregnant and support person participants were sent the same videos on the same schedule.

Participants in the control group were sent links to American College of Obstetricians and Gynecologists (ACOG) patient education webpages at enrollment. The links were sent once at enrollment but were accessible throughout the study and could be resent upon request. The webpages included answers to frequently asked questions on preterm birth, cesarean sections, and a partner’s guide to pregnancy.

At enrollment, participants completed items on sociodemographic information and their living situation. Patient-Reported Outcomes Measurement Information System (PROMIS) Anxiety (15) Computerized Adaptive Testing (CAT) assessed general anxiety over the past seven days. The PROMIS measures are converted to a normed T-score, with 50 (SD 10) representing the US general population average.

Follow-up assessments were administered at 25, 30, and 34 weeks GA if pregnancy continued. Pregnant and support person participants received the same follow-up assessments. The assessments were delivered through a REDCap link sent by text message or email, depending on the participant’s preference. Follow-up assessments included the PROMIS Anxiety CAT and the Parent Prematurity Knowledge Questionnaire (PPKQ). The PPKQ tests knowledge of long-term outcomes, lowest GA needed for survival, treatment options, and factors influencing outcomes (12). The assessment contained 38 items at week 25, 35 items at week 30, and 30 items at week 34. The percentage of correct responses was automatically scored in REDCap for each participant. One item of the PPKQ tests knowledge of unmarried fathers’ legal standing to make treatment decisions at birth. Of note, all participants were receiving care in Wisconsin where this legality is applicable (10). Questionnaires at 30 and 34 weeks included an item for pregnant mothers to self-report activities undertaken to prepare for preterm birth, including if they had decided to breastfeed. Participants were given $20 for each completed assessment.

P3 software tracked use of each video by each participant to assess use of the program.

The software collected data on the videos sent, the links that the participants clicked on, the videos played, and the time played in each viewing.

To allow investigation of the effect of father participation in prenatal education and reduce confounders related to father’s relationship with the mother, we excluded from analysis the mothers and fathers who were not living together at the time of enrollment. Participants not responding to questionnaires at the 25, 30 or 34 weeks GA timepoints (Figure 1) were not included in the analysis for that timepoint. However, as participants could respond to some but not all questionnaires, their results were included in analysis for timepoints when they did respond. Means were used to summarize continuous variables while counts and percentages were used for categorical variables. Each estimate is accompanied by the 95% CI.

**Figure 1.**
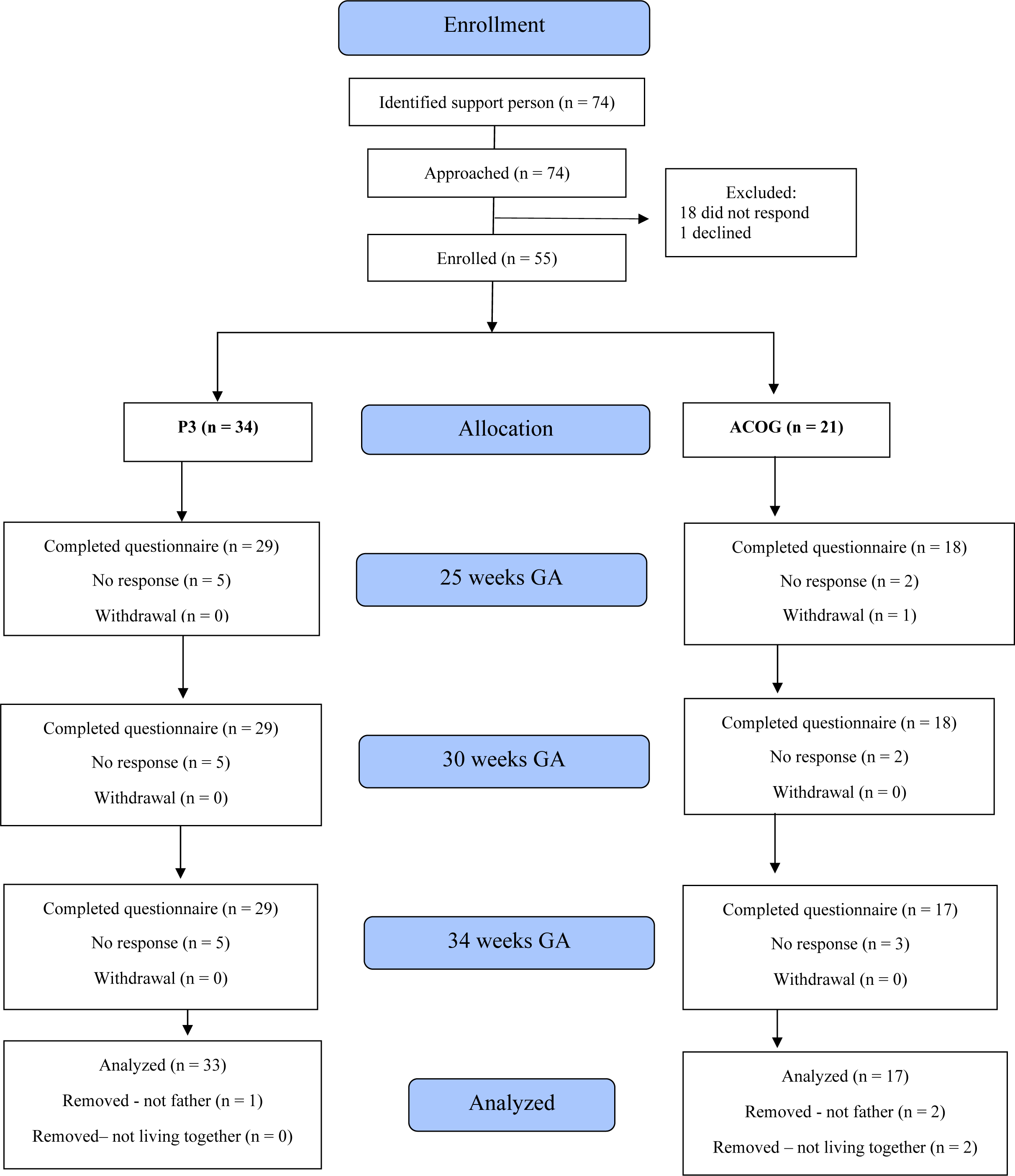
CONSORT flow diagram of support person participants.

## 3. Results

120 pregnant participants enrolled; 3 of the 120 pregnant mothers participated with a support person other than the father of the baby (2 with their mother [the baby’s grandmother] and 1 with their same-sex partner). These participant pairs were excluded from this analysis to avoid potential heterogeneity by support person type. Of the 104 mothers who reported living with the father of the baby, 66 (63%) provided contact information for the father and 50 (76%) of the invited fathers enrolled (Figure 1). The total sample included in this analysis therefore was 50 mother-father pairs living and participating together and 54 mothers living with the father but participating alone.

Fathers’ demographics did not differ by study group (Table 1). However, more fathers participated in the P3 group (n=33) than the control group (n=17), due to pre-randomization differences in provision of support person contact information by the mother (70% vs 47%) as well as differences in support person participation rates post-randomization (81% vs 68%).

**Table 1.** Participating fathers’ demographic information by study group.

| **Fathers** | **Total (N=50)** | **P3 (N=33)** | **Control (N=17)** |
| --- | --- | --- | --- |
| **Race,<sup>a,b</sup> n (%)** |  |  |  |
| Asian | 3 (6) | 2 (6) | 1 (6) |
| Black | 7 (14) | 5 (15) | 2 (12) |
| White | 41 (82) | 26 (79) | 15 (88) |
| Other<sup>c</sup> | 2 (4) | 1 (3) | 1 (6) |
| **Ethnicity,<sup>b</sup> n (%)** |  |  |  |
| Hispanic or Latino | 7 (14) | 6 (18) | 1 (6) |
| Not Hispanic or Latino | 43 (86) | 27 (82) | 16 (94) |
| **Education level, n (%)** |  |  |  |
| High school diploma | 6 (12) | 4 (12) | 2 (12) |
| Some college | 19 (38) | 16 (49) | 3 (18) |
| 4-year degree | 16 (32) | 10 (30) | 6 (35) |
| Graduate degree | 9 (18) | 3 (9) | 6 (35) |
| **Age, n (%)** |  |  |  |
| 20-24 | 3 (6) | 2 (7) | 1 (6) |
| 25-34 | 20 (42) | 12 (39) | 8 (47) |
| 35+ | 25 (52) | 17 (55) | 8 (47) |
| Missing | 2 | 2 | 0 |
| **Marital status, n (%)** |  |  |  |
| Married | 40 (80) | 25 (76) | 15 (88) |
| Not married | 10 (20) | 8 (24) | 2 (12) |

<sup>a</sup> Race percentages may not add up to 100%, as participants could select more than one race.
<sup>b</sup> Race and ethnicity were self-reported using National Institutes of Health prespecified categories.
<sup>c</sup> 'Other' included: Native Hawaiian, participant self-reported 'other', and participant preferred not to say.

Of the mothers who lived with the father of the baby, characteristics by participation with the father or alone can be found in Table 2.

**Table 2.** Characteristics of pregnant mothers who live with the father of the baby, by study group and whether they participated with the father of the baby or participated alone.

| Characteristics of pregnant mothers | Control group |  | p-value | P3 group |  | p-value |
| --- | --- | --- | --- | --- | --- | --- |
|  | Participating with fathers<br>N=17 | Participating alone<br>N=33 |  | Participating with fathers<br>N = 33 | Participating alone<br>N = 21 |  |
| Race,<sup>a,b</sup> n (%) |  |  | 0.43 |  |  | 0.15 |
| Asian | 1 (6) | 3 (9) |  | 0 | 1 (5) |  |
| Black | 1 (6) | 7 (21) |  | 3 (9) | 5 (24) |  |
| White | 15 (88) | 22 (67) |  | 28 (85) | 15 (71) |  |
| Other<sup>c</sup> | 0 | 1 (3) |  | 2 (6) | 0 |  |
| Ethnicity,<sup>b</sup> n (%) |  |  | 0.65 |  |  | 0.65 |
| Hispanic or Latino | 1 (6) | 4 (12) |  | 3 (9) | 1 (5) |  |
| Not Hispanic or Latino | 16 (94) | 29 (88) |  | 30 (91) | 20 (95) |  |
| Education, n (%) |  |  | 0.026 |  |  | 0.65 |
| High school or less | 1 (6) | 6 (18) |  | 3 (9) | 3 (14) |  |
| Some college | 2 (12) | 12 (36) |  | 10 (30) | 7 (33) |  |
| 4-year degree | 10 (59) | 5 (15) |  | 9 (27) | 8 (38) |  |
| Graduate degree | 4 (24) | 10 (30) |  | 11 (33) | 3 (14) |  |
| Age, n (%) |  |  | 0.43 |  |  | 1.0 |
| 20-24 | 1 (6) | 3 (9) |  | 3 (9) | 2 (10) |  |
| 25-34 | 8 (47) | 21 (64) |  | 22 (67) | 14 (67) |  |
| 35+ | 8 (47) | 9 (27) |  | 8 (24) | 5 (24) |  |
| Marital status, n (%) |  |  | 0.29 |  |  | 0.28 |
| Married | 15 (88) | 24 (73) |  | 25 (76) | 13 (62) |  |
| Not married | 2 (12) | 9 (27) |  | 8 (24) | 8 (38) |  |

<sup>a</sup> Race percentages may not add up to 100%, as participants could select more than one race.
<sup>b</sup> Race and ethnicity were self-reported using National Institutes of Health prespecified categories.
<sup>c</sup> ‘Other’ included: American Indian and participant self-reported ‘other’.

In the P3 group, the mothers who participated with the father had higher overall mean knowledge scores than those who participated alone at 25 weeks GA, 85% correct vs.76% (difference 9.2, 95% CI: 0.8, 17.5), at 30 weeks GA (though this did not reach significance), 87% correct vs. 82% (difference 5.1, 95% CI: -0.4, 10.6), and at 34 weeks GA, 90% correct vs. 84% (difference 6.4, 95% CI: 2.0, 10.8), Figure 2. In the control group, the knowledge scores of mothers who participated with the father were not significantly different than those who participated alone at 25 weeks GA, 67% correct vs. 60% (difference 6.8, 95% CI: -0.5, 14.1), 30 weeks GA, 74% correct vs. 68% (difference 5.6, 95% CI: -2.7, 13.9), or 34 weeks GA, 77% correct vs. 72% (difference 5.0, 95% CI: -1.9, 12.0), Figure 1 in supplement.

**Figure 2.**
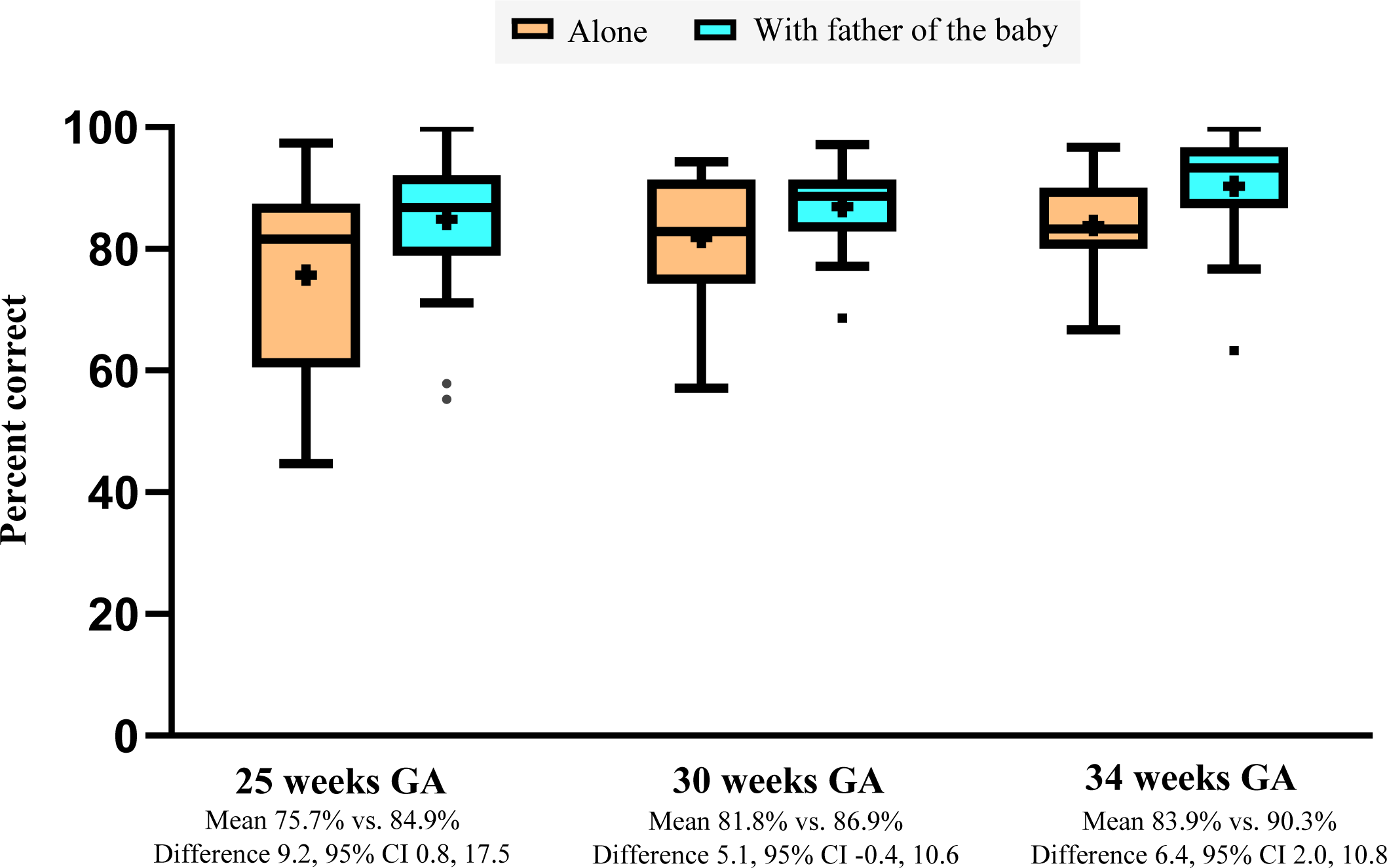
Overall knowledge scores at each time point among mothers in the P3 group by whether they participated alone or with the father of the baby. Mean scores indicated by ‘+’.

In the P3 group, the pregnant mothers who participated with the father watched an average of 64% of the videos (95% CI: 53.0, 74.9); mothers who participated alone watched an average of 48% (95% CI: 31.7, 65.6). There was a positive, moderate correlation between mothers’ viewing and fathers’ viewing, r=0.42. For every additional 10% of videos watched by the father, mother’s percentage of videos watched increased by 4% (standard error, SE=1.6%).

Fathers in the P3 group watched a mean (SD) 41% (32.1) of the videos sent (Figure 2 in supplement). Mean knowledge scores of fathers in the P3 group were not statistically higher than the scores of fathers in the control group: at 25 weeks GA, 77% correct vs. 68% (difference 8.4, 95% CI: -0.1, 16.9), 30 weeks GA, 79% correct vs. 73% (difference 6.1, 95% CI: -0.4, 12.7), and 34 weeks GA, 80% correct vs. 69% (difference 10.8, 95% CI: -2.2, 23.9), Figure 3.

**Figure 3.**
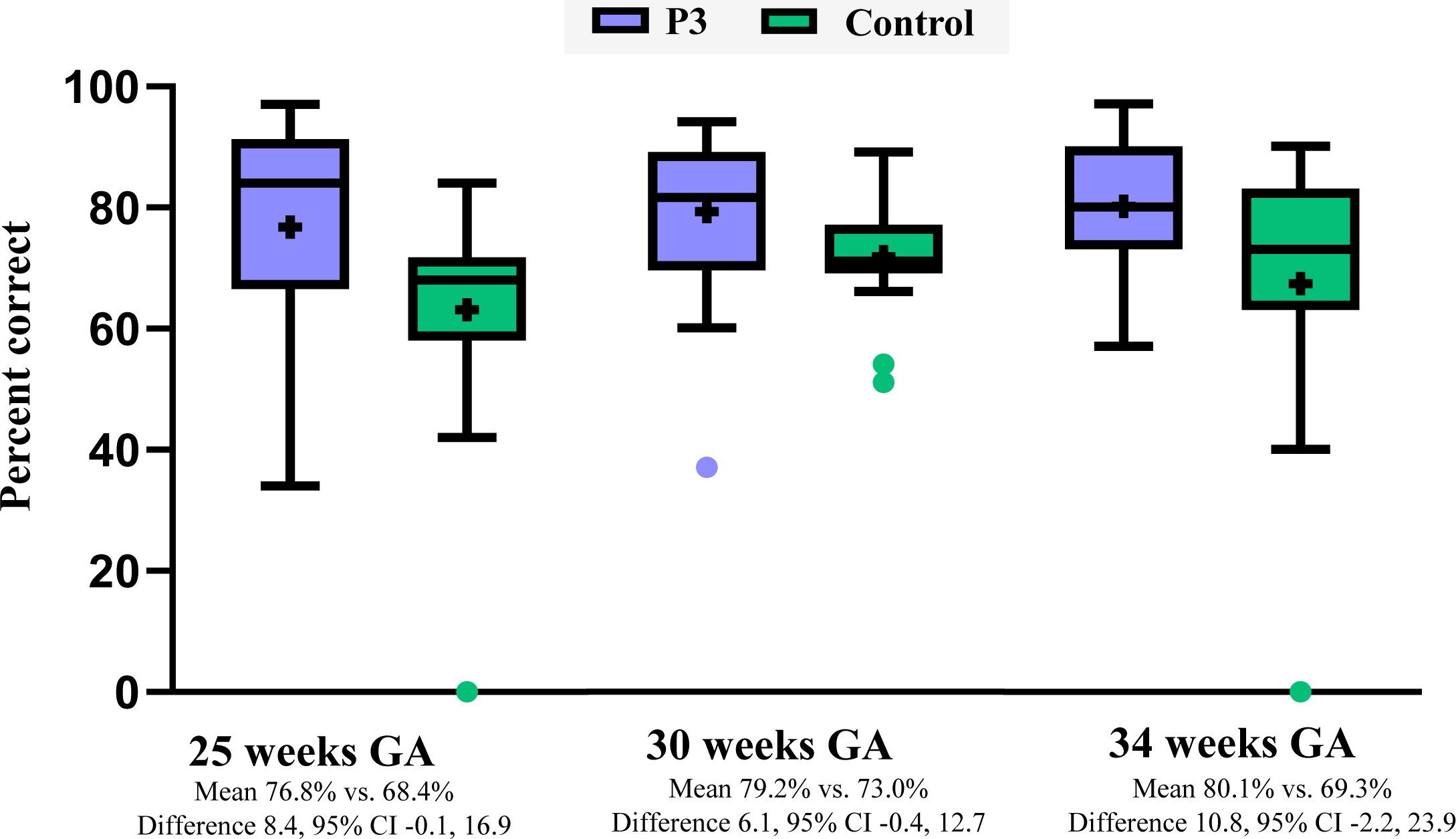
Participating fathers’ knowledge scores at each follow up, by study group. Mean scores indicated by ‘+’.

Fathers receiving the P3 education reported greater preparedness than the control fathers for making a neonatal resuscitation decision at 25 weeks GA, 77 out of 100 possible points vs. 59 (difference 18.3, 95% CI: 1.1, 35.4). The difference between study groups was not significant at 30 weeks for a birth hospital decision, 69 out of 100 vs. 62 (difference 7.9, 95% CI: -4.1, 19.9) or 34 weeks for a breastfeeding decision, 70 out of 100 vs. 63 (difference 6.2, 95% CI: -11.2, 23.6).

There was no change in fathers’ anxiety levels from enrollment for the P3 or control groups at 25 weeks GA (P3: difference -0.6, 95% CI: -2.4, 1.3; control: difference 0.7, 95% CI: - 2.2, 3.5), 30 weeks GA (P3: difference -0.6, 95% CI: -3.0, 1.8; control: difference 1.3, 95% CI: - 1.5, 4.1), or 34 weeks GA (P3: difference 1.3, 95% CI: -1.6, 4.1; control: difference 0.9, 95% CI: -3.4, 5.2), Figure 3 in supplement.

Across both study groups, among pregnant mothers living with the baby’s father, 45/47 (96%) of mothers participating with the father reported at 30 weeks GA having decided to breastfeed, compared to 32/45 (71%) of mothers participating alone. At 34 weeks, 43/48 (90%) of mothers participating with the father reported having decided to breastfeed, compared to 37/43 (86%) participating alone.

Despite its relevance for them, unmarried parents (n=37) were no more likely than married parents (n=117) at 25 weeks GA to know unmarried fathers’ legal standing at birth to make neonatal treatment decisions, 61% correct vs. 59% (difference 2.4, 95% CI -17.0, 21.8). However, parents who watched the P3 video conveying this information about unmarried fathers’ legal standing were more likely to answer these knowledge questions correctly at 25 weeks GA, 84% vs. 41% correct (difference 43, 95% CI 29, 58), with similar results at 30 weeks GA and 34 weeks GA (Figure 4).

**Figure 4.**
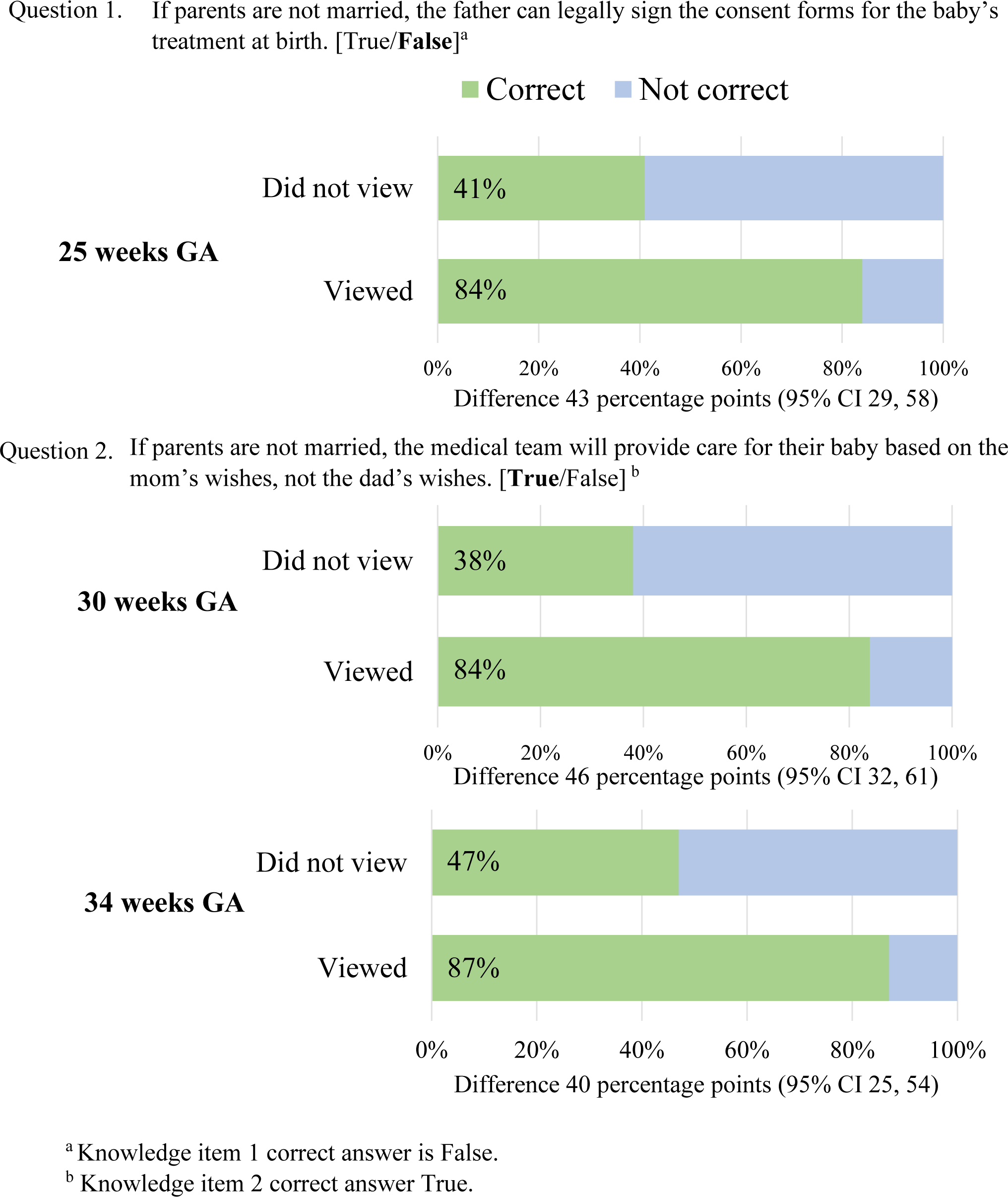
Knowledge items on unmarried fathers’ legal standing for neonatal treatment decision making, by viewing of the relevant video.

## 4. Discussion and Conclusion

### 4.1 Discussion

Among cohabitating opposite-sex couples with high-risk pregnancies, fathers’ participation enhanced maternal learning, as mothers who participated in the P3 program with the baby’s father had greater knowledge scores and greater intention to breastfeed than mothers whose partner did not participate. Additionally, parental awareness of unmarried fathers’ legal standing in Wisconsin for treatment decisions was greatly improved by viewing the associated P3 video.

Participating fathers engaged in learning from the P3 program without an increase in anxiety. While the mean of 41% of videos viewed is less than half the videos sent, that corresponds to about 50 minutes of educational videos. Given over one third of fathers do not attend any prenatal visits (16), each of which must cover a wide range of information in about 15 minutes (17), this is more exposure to premature birth information than most fathers likely receive from clinic visits. While there was no statistically significant difference in knowledge scores between P3 group and control group fathers, the trend toward greater knowledge in the P3 program (difference at 25 weeks GA, 8.4, 95% CI: -0.1, 16.9) suggests that this may be due to a type 2 statistical error. Additionally, the differing participation rates post-randomization (81% enrolling in P3 program vs. 68% in control) may indicate that fathers are more interested in learning about premature birth with the P3 program than through traditional methods such as patient education webpages.

The finding that mothers participating with fathers of the baby typically had more preterm birth knowledge may result from several sources. We speculate that their greater knowledge may be related to mothers participating with fathers in the P3 group watching more videos than those participating alone. Alternatively, as we previously reported in Flynn et al. (12), P3 mothers reported having more discussions with their partner about preterm birth than the control group; as the father was receiving the same video content, these discussions may have been more informed and reinforced the learning material. However, we cannot rule out the possibility that despite similar living situations, fathers’ baseline involvement may be higher among couples who participated together than when the mother participated alone. While in the control group, fathers’ participation differed by mothers’ educational levels (Table 2), the P3 group, which demonstrated the difference in knowledge, did not have demographic differences. Regardless, these results are strengthened by our exclusion of mothers who do not live with the father of the baby in the analysis. Much past work on paternal involvement measures fathers’ involvement by if his name is recorded on the birth certificate (2,18), but there is a broad spectrum of a father’s potential level of engagement in their child’s life. While engaging fathers who are unknown or not available would be beyond the scope of an educational intervention, many fathers are emotionally invested in the pregnancy (19,20) but not offered equal access to informational resources as mothers. Our finding that within similar levels of partnership (i.e., living together), fathers’ participation was associated with improved maternal knowledge is in line with previous studies demonstrating that paternal involvement is associated with mothers’ healthier prenatal behaviors (18) and demonstrates the potential power of fathers in pregnancy.

In many states, including Wisconsin where participants were receiving care, fathers are not legally considered parents at birth unless they are married to the baby’s mother (9,10).

Voluntary Paternity Acknowledgement forms have improved the rates of establishing legal parenthood (21), but they cannot be filed before birth and are not effective until after the filing, days to weeks later. During this interim period, only the mother can legally consent to treatments. In this study, responses to items on unmarried fathers’ legal ability to make treatment choices at birth demonstrate that even among unmarried parents, this legality is not common knowledge. Fathers in this situation may feel disempowered (22); this can become even more difficult in cases where the mother is incapacitated, as many were due to COVID-19 (23). In such cases, decision making for the baby may legally reside with persons who are not parents of the child (10). With awareness of this legality, parents would have the opportunity to have conversations about these situations, document their preferences, and share that information with family if appropriate.

This study is limited by its small sample size, which precluded analysis of other types of support person relationships (e.g., same sex partners, family members). While these support persons deserve investigation, aggregating their data would have missed potential nuances by support person role. An adequately powered study is also needed to draw more definitive conclusions about fathers’ results. Finally, our recruitment relied on mothers as gatekeepers to contacting the fathers for enrollment, which may have limited fathers’ opportunities to participate. However, because study inclusion is dependent on mothers’ health conditions as risk factors (e.g., chronic hypertension), independently recruiting these fathers would not have been feasible in this model.

### 4.2 Conclusion

Among opposite-sex couples cohabitating, the mothers who participated in the P3 program with the baby’s father had more preterm birth knowledge and more intention to breastfeed.

### 4.3 Practice Implications

Fathers face many barriers to information on pregnancy care, including for pregnancies with risk factors for preterm birth. Consequently, fathers are unprepared for preterm birth and mothers are burdened by not having an equally informed partner. The P3 program’s ability to overcome clinic-based barriers and provide preterm birth information to fathers may benefit families with high-risk pregnancies.

## Data Availability

Our research team will consider requests for re-use of scientific data from scientists of like-minded interests, and these will be prepared through MCW. No data sharing will take place until a signed data sharing agreement is in place, necessary costs of sharing provided, and all site PIs are aware and approve.

